# A disorder of consciousness rather than complete locked-in may be the final stage of ALS

**DOI:** 10.1101/2024.06.21.24307994

**Authors:** F. Gobert, I. Merida, E. Maby, P. Seguin, J. Jung, D. Morlet, N. André-Obadia, F. Dailler, Ch. Berthomier, A. Otman, D. Le Bars, Ch. Scheiber, A. Hammers, E. Bernard, N. Costes, R. Bouet, J. Mattout

## Abstract

The end-stage of amyotrophic lateral sclerosis [ALS] is presumed to be a complete Locked-In Syndrome [cLIS], assuming an internally preserved consciousness that would not be accessible anymore from the outside. However, whether consciousness persists at this stage of ALS remains to be demonstrated. Shifting the perspective from cLIS (presupposed consciousness) to Cognitive Motor Dissociation (to-be-demonstrated consciousness), we attempted to demonstrate consciousness and communication with two cLIS-ALS patients using a multimodal awareness assessment battery. It involved complete neurophysiological assessments, passive and active auditory oddball paradigm (Subject-Own-Name/P300), an auditory-based Brain-Computer-Interface [BCI] and activation-task imaging using functional MRI or [^15^O]H2O PET. Wakefulness (long-term EEG), brain morphology (CT or MRI scans) and resting brain metabolism ([^18^F]fluoro-deoxy-glucose PET) were used to describe the underlying cLIS brain function.

While Patient 1 could initially follow simple commands, he failed twice to control the BCI. At follow-up, he showed no more evidence of command following and his oddball (Own Name - P300) cognitive responses has disappeared. At his unique evaluation, Patient 2 was neither able to follow simple commands nor to control the BCI.

Both patients had altered wakefulness, brain atrophy, and a global cortico-sub-cortical hypometabolism pattern compatible with a disorder of consciousness, regarded as an extreme form of an ALS-associated fronto-temporal dementia.

While it is not possible to firmly demonstrate the absence of awareness, each independent measure concurred with suggesting that a “degenerative disorder of consciousness” rather than a cLIS might be the final stage of ALS. In future cases, this dramatic cognitive decline should be anticipated before communication disappears to enable precise advance directives regarding end-of-life issues in case complete – and neurophysiologically confirmed – unresponsiveness occurs.

Altogether, the neuroimaging features distinguishing the mechanisms in this rare condition is a significant milestone to understand end-stage ALS. The present clinical study calls for further exploration of this terminal stage to determine the prevalence of this profile in whom communication seems hopeless.

## Introduction

Amyotrophic lateral sclerosis [ALS] is a neurodegenerative disorder characterized by a progressive motor weakness with muscle atrophy leading to tetraplegia and respiratory distress, with death occurring 2 to 3 years from onset. ALS is associated with frontotemporal dementia (FTD) for 15% of patients, but cognitive and behavioural impairments occur in up to 80% by end-stage disease^1^. Extra-motor extension of ALS related-abnormalities has previously been described for brain metabolism^2, 3^, neurotransmitter function^4, 5^ or perfusion^6^ at the population level but are rarely prescribed for diagnosis purposes at the individual level. Morphological biomarkers of initial ALS without cognitive impairment are focused on corticospinal tract involvement^7^. Abnormal encephalic [^18^F]fluoro-deoxy-glucose [FDG] Positron Emission Tomography [PET] has been proposed as a fairly accurate and early diagnosis tool in ALS^8, 9^, while cortical morphometry is normal at the early stage^10^. In case of early cognitive impairment, frontal FDG-PET hypometabolism could be associated with motor and somatosensory cortical atrophy, in line with the diagnosis of ALS-FTD^2^. In case of ALS without cognitive or behavioural symptoms, the atrophy could be rather observed in the brainstem^10^.

As riluzole and non-invasive ventilation are^11^ currently the only therapies extending survival^12^, symptomatic treatments are focused on improving the quality of life and delaying the loss of autonomy^13^. Therefore, continuing an eye-based communication (saccades or blink) after speech disappearance is critical. Saccades are classically spared in early ALS^14^ but subclinical saccadic disorders have been detected as an early sign of a frontal decline^15^. Beyond the point of saccade-based communication disappearance, the functional state associated with the ultimate stages of ALS has been poorly investigated but clinical reports of late symptoms suggested a large diffusion of the neurodegenerative pathology involving sphincterian and complex oculomotor disorders^16^. Histological data confirmed widespread loss of pyramidal neurons in frontal cortex and anterior cingulate areas^17^. In most cases, this last stage is not reached because tracheotomy is not proposed, refused, or an unexpected lethal complication occurs earlier. If chosen, tracheostomy, by extending survival to an average 7 years^18^, might also increase the probability of reaching these extreme stages of the disease^19^. Proposing and performing tracheostomy is highly dependent on the healthcare system and cultural background, varying from 5% in Europe^20^ to 30% in Japan^21^.

In rare instances, the final cognitive status cannot be behaviourally measured because of a complete motor failure that forbids any inter-individual communication. At this point, the clinical picture in this degenerative process resembles an entity which in the post-acute brain literature has been described as “Locked-In Syndrome”, presenting at least minimal facial/ocular movements after severe brainstem vascular injury^22^. The concept of functional^23^ or complete^24^ Locked-In Syndrome [cLIS] (that could be included in the recent concept of “cognitive-motor dissociation”^25, 26^) was then introduced. It hypothesizes that paraclinical tools could assess cognitive or consciousness contents in spite of the inability to behaviourally demonstrate them^26, 27^. In the ALS context, trying to communicate with cLIS patients by non-motor ways is of utmost importance for practical and ethical reasons^28^: to adapt treatment to pain and to confirm the patients’ will to carry on supportive care in the absence of daily communication. This motivated the development of brain-computer interfaces dedicated to the restoration of these patients’ communication^29^. However, as BCI tools have not been proven useful in the cLIS context so far^30^, several attempts were performed to define whether some consciousness dimensions could be impaired in the ALS-related cLIS context but neither polysomnographic sleep-wake patterns^31^, EEG spectrum^32^ not EEG complexity^30^ were found sufficiently altered so far to conclude definitely on the existence of “disorders of consciousness” [DOC]. However, simple brain responses usually tested in DOC (Somatosensory Evoked Potentials^33^) were found absent or altered in other cohorts, indicating the heterogeneity of the end-stage ALS population and the possibility of more severe stages of the neurodegenerative process.

In the present study, we adapted a “multimodal awareness assessment battery” [MAAB] to a neurodegenerative context, after a cLIS-like progressive loss of saccade control among two ALS patients with prolonged survival after their tracheotomy. For a comprehensive phenotypic description of their consciousness level, wakefulness fluctuations and brain metabolism were investigated as well.

## Results

### Population description

At the date of the first recording, Patient 1 (male, 56-60 years old) and Patient 2 (male, 70-75 years old) presented a 5- and 8-year evolving ALS, respectively. Communication was lost 3 and 9 months earlier, respectively (Figure 1).

Patient 1 was able to communicate using an eye-tracking system until Year 4. Saccades slowed down progressively, but eye movements could still be recognised by his relatives. In March of Year 5, the saccadic amplitude was too weak to be reliably perceived by a trained neurologist. The final complete assessment (T2) took place 13 months after the loss of communication by saccadic eye movements.

Patient 2 was able to choose from a list until Year 6 and to have a reliable “Yes-No” code until the end of Year 7. Responding to simple commands by saccades was still possible until January of Year 8 after strong verbal stimulations by a speech therapist.

In both cases, after having excluded confounding local factors of saccade-based communication inability by an ophthalmologic examination, a cLIS was hypothesised despite their cognitive ability to communicate being unknown.

The behavioural classification of consciousness based on motor responses (i.e. Coma Recovery Scale-Revised [CRS-R]^34^) was inconclusive to demonstrate any cortical function for both patients (CRS-R score = 2, with fixed eye-opening without stimulation related to a bilateral facial palsy). Patient 1 presented no eye-movement at all, but Patient 2 had a spontaneous and transient gaze deviation to the right that could be regarded as a pathological ping-pong gaze with a limited amplitude on the left^35^. Neither significant voluntary control of movement nor visual-guided saccades was possible to the right.

As the patient management was modified to answer a clinically relevant question (communication ability by paraclinical tools) and not for research purposes, this individualised analysis was conducted in accordance with the law on data protection (version n°2004-801, 6 August 2004). The results of neurophysiological and imaging explorations were given to the physician in charge of patients’ treatment. In addition, informed consents to transfer the patients to the teaching hospital for the MAAB were obtained before starting the exploration from legal representatives.

Authorization was obtained from the ethics committee of the Hospices Civils de Lyon (Comité Scientifique et Éthique des Hospices Civils de LYON CSE-HCL – IRB 00013204; Pr Cyrille Confavreux; approval N. 24-310).

### Absence of control of an auditory BCI (Table 1)

For both sessions of Patient 1 and the single session of Patient 2, the online test accuracies were at chance level (Table 1).

In contrast, cross-validation accuracies within sessions were above chance level (see Table 1), which typically reflect overfitting (*Table 1*).

### A disappearance of responsiveness confirmed by passive Evoked Potentials and active processes (Figure 2)

For Patient 1, at T1 the N100 response and the MMN response to duration deviant and the novelty P3 response to the patient’s own name were clearly identified. At T2, the novelty P3 was not identified but the N100 and MMN response to deviants persisted. For Patient 2, the neurophysiological pattern was comparable to Patient 1 at T2 (excepted the left peripheral deafness leading to adapt the BCI protocol) without passive Subject Own Name [SON]-P300 responses.

For Patient 1 at T1, the active paradigm showed a significant and diffuse P3b response to targets in the focused-attention condition (“Count”). These components were not found when the attention was diverted from the auditory stimuli by the mental imagery task. A significant and diffuse N2 response could be observed when comparing the responses to deviants in the focused-attention and diverted-attention conditions (“Focused *vs* Diverted”). No specific responses could be detected for Patient 1 at T2.

For Patient 2, no response was observed in the “Count” condition. A significant P3 response was observed for the “Focused *vs* Diverted” condition but it was rejected as inconsistent during the clinical review process as it appeared on a single derivation (Pz) and with a late latency (750 ms).

### An alteration of wakefulness confirmed by long-term EEG (Figure 3)

For Patient 1 at T1, the standard EEG showed a physiological background activity with Alpha rhythm in posterior regions with spontaneous fluctuations. Background EEG activity was reactive to external stimulations such as light, touch, and sound (Fig SOM-1). At T2, EEG suggested fluctuations of vigilance with an alternating pattern with spontaneous diffuse theta activity alternating with delta activity on a long-term recording. A prolonged period of “abnormal wake” was distinguished from two periods with increased Delta power during the night, suggesting the persistence of slow-wave sleep. This wakefulness pattern was different from the normal arousal pattern of a healthy subject and from the spectral analysis on 20 minutes at T1. The sleep classification by ASEEGA algorithm^36, 37, 38^ (Physip®) was compatible with N2-N3 periods during the period of increased Delta power but the mixed Theta-Alpha periods were instable and alternatively classified as N1, REM sleep or wake.

For Patient 2, brain activity was depressed but labile with few physiological sleep figures. A slight reactivity of background EEG rhythm to sound and touch remained present (Fig SOM-1). The sleep/wake architecture was disrupted despite sleep-like patterns occurring preferentially during the night (e.g. numerous N2 and N3 periods between 10PM to 6AM). However, N3 occurred also during the day (e.g. between 4:45 PM to 6PM) and prolonged arousal periods were observed during the night (e.g. around midnight).

### Brain imaging provided no further argument of active processing but could explain the overall pattern by severe atrophy and hypometabolism

The morphological scans (Fig 4, top) demonstrated substantial brain atrophy, either in the frontal and temporal lobes (Patient 1) or more diffusely (Patient 2).

FDG-PET hypometabolism (Fig 4, middle) was global (including in the thalamus and caudate nucleus) for both patients. According to the analysis of variance corrected for age and global metabolism, no cortical region showed relative hypometabolism or hypermetabolism. The significant clusters of relative hypometabolism were rejected because of their extra-cerebral location in Figure SOM-2, top: left lateral sulcus for Patient 1, lateral ventricles for Patient 2). The significant clusters of relative hypermetabolism (Figure SOM-2, bottom) were in both cases in the periventricular white matter. This was related to a scaling issue due to the global mean correction. It created a statistical artefact as white matter brain areas were relatively less hypometabolic for both patients compared to the grey matter (mostly involved in the global hypometabolism). In other terms, the hypometabolism of white matter (whose absolute reduction of metabolism had a lower amplitude that the grey matter absolute reduction of metabolism) appeared as pseudo-hypermetabolic. This indirectly reinforced the argument for the cortical hypometabolism.

These combined analyses indicated that no voxel had a preserved metabolism within the fronto-parietal cortex because: this cortex was mostly involved in the global hypometabolism (Fig 4, middle); neither global (Fig 4, bottom) nor relative (Figure SOM-2, bottom) hypermetabolism were found in this region.

Neither cerebral blood flow changes in [^15^O]H2O-PET/CT (Patient 1 – T2) nor BOLD modulation in functional MRI [fMRI] (Patient 2) showed any difference in parahippocampal areas during the mental imagery task (spatial navigation^27, 39^) when comparing active and passive scans (data not shown).

## Discussion

Several specialised centres have proposed BCI to communicate with patients in end-stage ALS. In most of the cases, they did not achieve consistent on-line communication^30, 40^ but in some cases, succeeded^41^. Technical^40^ and theoretical^42^ limitations have been emphasised so far to explain failures, putting the discussion concerning the impact of pathology on the back burner^40^. However, no neuroimaging data were previously proposed to explore the central evolution during this final global neurodegenerative condition. Our multimodal explorations shed some light on this alternative explanation: BCI failed in some late-ALS patients – previously communicating by saccades – not only because eye-movements vanish, attentional processes decrease or the algorithmic interfaces were sub-optimal: in some cases, the most probable cause could be an evolving degenerative DOC. This state could be considered as the endpoint of the ALS-FTD continuum.

We described herein a complete loss of communication and even of response to command (excepted in a transitional recording T1 for Patient 1), as assessed with multiple paraclinical tests including a BCI, in two ALS patients who both had demonstrated communication more than four years after onset of tetraplegia and tracheostomy. Both lost behavioural functional communication less than one year before the first neurophysiological attempt to assess their cognitive residual abilities. Patient 1 demonstrated a neurophysiological response to simple command for at least three months after the last saccade-based behavioural communication. We decided to try a non-invasive BCI in order to restore communication. The auditory modality was proposed as oculomotor impairment is known to prevent the use of visual BCI^16, 43, 44^. But there are now multiple arguments to support that the absence of control of the BCI was due to the intrinsic evolution of the causal disease, as previously suspected in the literature^42, 45^.

In a previous study^24^, electro-corticography (ECoG) was used for four sessions of event-related potentials (semantic and auditory stimulus) during six months. The results remained stable before the last communication using motor contact. Three months after the patient fulfilled cLIS criteria, P300 responses were no longer detected, concomitantly with a persisting MMN and an increased power of low frequency bands. This last neurological state was not discussed as an end-stage ALS or dementia but rather as the consequence of an intercurrent infection. Similarly, another study focusing on semantic cognitive process in a single patient with a cLIS found a progressive loss of responses to oddball and semantic paradigm but neither classic MMN nor imaging were performed to further describe this phenotype^46^. More generally, several electrophysiological features seem to be altered in advanced stages of ALS^41, 47, 48^.

Neurophysiological signs of awareness were still observed three months after the last communication of Patient 1 (T1), as far as clear effects in the “Count” and “Focused *vs* Diverted” conditions were observed in the active auditory oddball paradigm. This patient might have been less cognitively impaired at T1 (before a final cognitive worsening at T2 when the SON-P300 disappeared as well) than the one from in the previous ECoG study^24^ and Patient 2 who presented an inconsistent response to the Act-Pass paradigm at his unique MAAB. One could argue that each of these cLIS candidates presented a particular dynamic of a common cognitive deterioration. In this hypothesis, the contingent dates of neurophysiological assessments would provide few screenshots of a shared continuum, ranging from a transient cLIS with attentional disorders (Patient 1 at T1) to an MCS-like status with difficulties to answer simple commands (Patient 2) and ultimately a more severe type of DOC, mimicking an Unresponsive Wakefulness Syndrome with abnormal EEG background rhythm associated to the complete loss of command following (Patient 1 at T2 and every other unresponsive patient from the litterature^24^).

### Absence of direct consciousness

Assuming that awareness can only be demonstrated by its presence when a reportable communication is confirmed, isolated negative BCI results were a necessary but not sufficient proof of awareness impairment. As the behavioural/motor gold standard was unavailable to confirm the presence or absence of awareness, we reinforced the methodological confidence by using alternative methods. Eventually, the MAAB was performed three times to test the following null hypothesis “the patient has no sign of awareness” using complementary levels of analysis. For Patient 1, we were initially able to reject this hypothesis in June of Year 5 (T1, before BCI testing) thanks to the patient’s ability to consistently follow a simple command during the Act-Pass session. For Patient 1 at T2 and for Patient 2, three consecutive negative tests (failed auditory BCI, absence of consistent responses to the Act-Pass paradigm, and failed activation-task imaging) did not allow to reject the null hypothesis. In other words, the presence of any reportable consciousness could not be proven any more at the individual level. At the general and theoretical level, it implied that we rejected the hypothesis stating that “end-stage ALS patients have only a behavioural communication issue” because DOC can occur at a final stage.

As neither neurophysiological nor imaging methods have been validated to diagnose the absence of awareness, the proposal of absence of awareness remained hypothetical and had to be cross-validated by complementary arguments confirming the impairment of basal brain functions supporting the global state of consciousness^49,50^.

Regarding the wakefulness dimension of consciousness^50^, EEG background activity for Patient 1 at T1 was compatible with a normal wake pattern, as previously described in typical cLIS patients^31^. In line with previous results^24^, a progressive slowing of EEG background rhythms was observed for Patient 1 at T2 and for Patient 2 during day-time EEG. Long-term EEG monitoring ruled out the hypothesis that wakefulness fluctuations (with sleep occurring unexpectedly during the day) could be solely responsible for the loss of all response to passive and active auditory paradigm: the sleep/wake architecture was abnormal but N2 and N3 occurred predominantly during the night for both patients and the “pseudo-wake” patterns predominating during the day were abnormal as well. Altogether, the EEG patterns were compatible with the continuum between sleep-like and a pathological (coma-like) state of wakefulness^51^ but as EEG was not continuously recorded during every passive or activation task, unexpected sleep or sleepiness period could not be formally ruled .

Concerning the assessment of cortical function required for consciousness^52^, the deteriorated neurophysiological responses were compatible with an altered processing of cognitive information^53^. An altered signal/noise ratio due to muscle artefacts was highly unlikely in ALS, given the depressed muscular tone. Recent EEG data confirmed the impairment of the EEG spectrum (shift towards slow waves at rest) in cLIS-ALS patients^32^, that was not related to an awareness issue by the authors.

Metabolic data gave strong arguments to integrate morphological and functional information into a reliable framework. In a comparative study between MRI voxel-based morphometry and FDG-PET in patients with ALS-FTD^54^, the hypometabolism was more widespread than the decrease of cortical thickness and better correlated to the clinical features. This earlier occurrence of decreased metabolism before the appearance of atrophy on morphological imaging may be explained by the relationship between the functional FDG-PET imaging and the synaptic activity itself. FDG is metabolised in mitochondria at the synaptic level and its uptake is correlated to the synaptophysin level, a marker of synaptic density^55^. Despite this physiological argument, confirming whether the metabolic impairment anticipates the extension of atrophy in ALS would require rigorous longitudinal studies.

Extrapolating from the global extension of hypometabolism and the related clinical severity^2^, we argue the present snapshot of ALS evolution describes a further step in ALS pathophysiology. Indeed, the extent of the “full-blown FTD” ^2^ hypometabolism was not comparable to the one observed in the fronto-parietal areas^56^,

^57^ among the present end-stage ALS patients, more in line with Unresponsive Wakefulness Syndrome patients, the most severe expression of chronic DOC. The involvement of extra-frontal areas could explain how a limited cognitive dysfunction evolves towards consciousness impairment, in line with both the global neuronal workspace hypothesis^58^ (fronto-parietal functional impairment) and the mesocircuit hypothesis^59^ (basal ganglia functional impairment).

Altogether, and contrary to other authors maintaining a segregation between the cLIS and DOC litteratures^30, 32, 33^, we hypothesise that late-stage cLIS-ALS can evolve through a FTD-ALS continuum to a progressive extinction of consciousness related to a multisystemic central involvement. It impairs wakefulness (as in the present polysomnographic results and in^31^), spectral and complexity EEG prerequisites of awareness (as in the present EEG results and in^30, 32^), command-following in different paradigms such as active oddball paradigm, activation task imaging and BCI adapted to the auditory stimulus (as in the present attempt and in^30, 33^), robust primary cortex responses (as in the present abnormal SEPs response with a reduced amplitude of the cortical response for Patient 2 and in^33^) and resting metabolism (as in the present unique demonstration). Two mechanistic hypotheses can be proposed to explain this final ALS stage.

First, a progressive “extinction of thought”^16^ can be discussed. It would be primarily due to the loss of afferent stimulus during cLIS, explaining why this extreme stage was proposed as an exclusion criterion to test a reliable communication device, based on a meta-analysis in various aetiologies (but mostly in ALS)^42^. In this hypothesis, the global hypometabolism and the subsequent atrophy would be a consequence of the cLIS rather caught out by a subsequent inability for any motor mental imaging.

Second, the dynamic of the cognitive decline could be compatible with the final stage of the ALS neurodegenerative process as previously described in other contexts such as Creutzfeldt-Jakob disease [CJD] in case reports^60,61^ or small series^62^. Despite the classic existence of neuro-cognitive impairment in ALS^63^, the respective speed of severe cognitive dysfunction (fast in CJD and slow in ALS) and respiratory failure (slow or absent in CJD and fast in ALS) implies that this final stage would be more probably observed among CJD cases. The brain metabolism of ALS-FTD might reach after years the functional level comparable to the one observed a few months after CJD onset. This hypothesis was also in line with the pathological features^19^ of cLIS showing the impairment of the mesocircuit^59^ and of the brainstem reticular formation^25^.

Of note, the present description does not allow causal inference to disentangle them.

### Limitations of the study

This description had several limitations. The main limitation is the small number of patients that could be included in this observatory study, due to the extreme rarity of such cases in a single ALS centre in France where most patients declined artificial ventilation, and by the necessity to avoid moving patients from other centres which would have provided them access to our complete awareness evaluation.

Therefore, these results should be confirmed in further studies and possibly with other forms of neurodegenerative diseases. Moreover, the pattern of diffuse hypometabolism could be compared to a PET database including conscious ALS and ALS-FTD patients. In the future, a longitudinal study should describe more ALS patients after the loss of oral communication and compare them to LIS or cLIS arising from different mechanisms and to severe FTD case in the absence of ALS. However, managing such a study seems difficult due to logistical issues and the rarity of such cases in a restricted area (as a combination of the ALS incidence and the willingness to survive in a cLIS condition).

## Conclusion

The comprehensive description of these two cases suggests that, for patients after a prolonged period of mechanical ventilation allowing survival beyond the natural history of ALS, the extreme evolution of an associated ALS-FTD pattern could induce a complete loss of communication which could resist to current paraclinical tools implemented to overcome the motor failure. Altogether, these patients might have reached a DOC stage instead of a functional LIS^23^. Thanks to a coherent corpus of arguments based on neurophysiological and imaging approaches, we proposed that such cases match more accurately to the description of a “degenerative DOC”. This point could have important psychological and ethical consequences for patients entering the final phase of ALS: waiting for BCI optimisation and success remains legitimate but this perspective appears as equivocal. Informing patients and caregivers that consciousness itself could be impaired at last is of utmost interest to plan state-of-the-art advances directives.

Ethical discussions in DOC after an acute brain injury usually face several uncertainties. In absence of explicit written patients’ will, what level of disability would have they considered as acceptable? The time course of ALS allows to address these questions in advance, when the patient keep his communication abilities. Another difference between ALS and other post-ictus recovering DOC is the dynamic of the disease (negative vs plausibly positive). The quite similar progressive loss of communication illustrated here (the delays between steps may vary) reduces the uncertainty concerning the prognosis: once a cLIS is observed and consciousness impairment is confirmed using a dedicated battery, no return to communication should be expected in absence of active treatment.

More theoretically, this study illustrates how much the diagnosis of disorders of consciousness is a conceptual challenge in the absence of any behavioural clue. However, this challenge could be overcome by convergent paraclinical arguments demonstrating the absence of awareness, as shown here. More importantly, the response can be reinforced by the absence of major prerequisites of awareness, such as physiological changes of wakefulness and a maintained resting metabolism.

## Materials and methods

### 1. Electrophysiological assessments

#### EEG recordings

EEG data were recorded according to a routine clinical protocol, using a Micromed**®** EEG recording system (11-electrode EEG for 20 min and 15-electrode monitoring system for 24h). EEG data were then interpreted by two trained neurophysiologists (NAO and FG). The long-term EEG was analysed using the ASEEGA algorithm of automatic sleep classification (CB, Physip®) using the Cz-Pz derivation and trained on normative data from healthy subjects^36^. It is illustrated by the data of a healthy subject.

#### Evoked potential protocol

Standard EP data were recorded i) for Patient 1 at T1 and for Patient 2 with a 7-electrode Micromed**®** EEG recording system (Fz, Cz, Pz, F3, F4, M1, M2) and ii) for Patient 1 at T2 with a 32-electrode BrainAmp**®** EEG system. The reference electrode was placed on the tip of the nose and the ground electrode on the forehead. One bipolar electro-oculogramm (EOG) derivation was recorded from 2 electrodes placed on the supra-orbital and infra-orbital ridges of the right eye. The signal was amplified (band-pass 0.3–100 Hz), digitized (sampling frequency 1000 Hz) and stored for off-line analysis.

Data from two auditory paradigms were obtained at each MAAB. The *passive oddball paradigm* was based on short duration deviant tones (occurrence frequency of 0.14) and included the patient’s own first name as a rare stimulus (“novel” with an occurrence frequency of 0.03). This protocol aimed at evidencing sensory auditory processing (N1 to standard tones), automatic change detection (mismatch negativity, MMN detected in the difference “deviants minus standards”) and attention orienting (novelty P3 in the response to novels). The patient was presented with one block of 2000 stimuli, including 280 deviants and 60 instances of his own name^64^. The *active-passive oddball paradigm* (Act-Pass) relied on frequency deviant tones (p = 0.15) and aimed at evidencing voluntary processes^65^. The auditory stimulation was presented in two successive conditions, the patient being first asked to actively divert his attention (using a mental imagery task) and then to actively focus his attention (counting the deviants). In each condition, the patient was presented with 3 blocks of 200 stimuli including 30 deviants, each block being preceded by recorded instructions. Voluntary processes were evidenced both by comparing responses to deviants and targets in the condition of focused-attention (“Count”) and by comparing responses to deviants in the focused-attention and in the diverted-attention conditions (“Focused *vs* Diverted”).

Wave detection was achieved in a two-step process including visual identification by a trained neurophysiologist^65^ and objective confirmation by means of randomization methods applied to the significant time-clusters. A one sample T-test was calculated between the two conditions (sample by sample; p-value < 0.05). A time-cluster was defined when many consecutive samples were significant. A cluster permutation^66^ was used to solve the multiple comparisons problem, incorporate biophysically motivated constraints in the test statistic and control the false alarm rate. According to this nonparametric method we build a permutation histogram based on 1000 random partitions of our two conditions and clusters were rejected if their statistics were above threshold (unilateral p-value < 0.05).

### 2. Auditory-based Brain Computer Interface

We developed a dedicated auditory Brain Computer Interface (BCI) based on spatial selective attention^67^ as standard visual BCI paradigms were not suitable in this case of complete oculomotor failure^68^. This auditory-based BCI was designed as a communication device to elicit a single reliable yes-or-no answer at a time: two interleaved trains of stimuli are presented and the BCI is driven by the difference in responses between all attended and all unattended stimuli.

We used spoken words pronounced by a synthesized male voice (“yes” and “no”) delivered in a pseudo-random order. This paradigm was benchmarked with 18 control subjects and 7 patients with severe motor disability. Seventeen out of 18 healthy subjects obtained online BCI accuracy above chance level. Results were more mitigated amongst patients as two patients with advanced ALS were at chance level, while the other two could control the BCI with perfect accuracy^68^. In contrast, the three patients with locked-in syndrome due to a brainstem injury could not control the BCI. A similar paradigm was developed and tested by Hill et al, 2014, reporting accuracies above chance level for both patients with ALS that tried the BCI^69^.

To answer a question, 20 standard stimuli (of 100 ms duration) and 3 deviant stimuli (of 150 ms duration) were delivered with a random Stimulus Onset Asynchrony between 400 ms and 700 ms. The patients had to orient their attention to the right side in order to answer “Yes” and to the left in order to answer “No”.

EEG referenced to the nose was recorded from 22 active electrodes (Acticap system-Brain Products, Germany) at positions Fp1, Fp2, F7, F3, Fz, F4, F8, FC5, FC1, FC2, FC6, C3, Cz, C4, CP5, CP1, CP2, CP6, P7, P3, P4, P8 following the extended 10-10 system placement^70^.

All parameters learned from the calibration session were subsequently applied during the test sessions. These parameters pertained to the feature selection and subsequent classification steps. The former consisted in spatial filters derived from the xDAWN algorithm^71^. Then a two-class Naive Bayes classifier was trained, based on the spatially filtered training data. More information on this classifier can be found in Seguin et al.^68^. At the end of calibration, a leave-one-out based cross-validation was used to evaluate the quality of this calibration. If cross-validation was above chance level, we performed the online testing of the BCI. On the contrary, if cross-validation did not exceed chance level, we started a new calibration. The course of each patient’s sessions is summarized in *Table 1*.

For the test session, the EEG stream was processed in real-time by a framework developed in Python: a band-pass filter between 0.5 and 20 Hz was applied, 500 ms long epochs were extracted to analyse the responses evoked by standard “yes” or “no” sounds.

Averaging was performed for each of these two conditions: standard “yes”, standard “no”. The Bayesian classifier computed the posterior probability for each class given the observed features (“yes” or “no” target stimuli versus “yes” or “no” non-target stimuli). Then the output of each classifier was optimally combined to obtain a final and unique posterior probability for each of the two classes (“yes” or “no”) (see the appendix of Seguin et al.^68^). Complementary offline analyses were performed to assess BCI performance based on different stimulus types (standard, deviant, yes, no).

Patient 1 underwent two BCI sessions one week apart (6 years after ALS onset, 10 months after the T1 initial MAAB, 2 months before the T2 complete MAAB). In the first one, patient 1 was asked to follow 20 instructions in the calibration session and 40 questions on basic general knowledge for the test session. For the second session, in an attempt to make the task more accessible, we decided to slow down the stimulation trains with a random Stimulus Onset Asynchrony between 600 ms and 900 ms.

Patient 2 underwent only one BCI session (8 years after ALS onset, and concomitantly to the other components of the MAAB) with an adapted (monaural) protocol because of a left peripheral deafness (as also described in Seguin et al.^68^ for one patient).

### 3. PET and fMRI Study

#### Acquisition of images

Three-dimensional PET scans were acquired on a PET/CT tomograph for Patient 1 (Biograph mCT/S 64 scanner) along with a morphologic CT scan to assess brain atrophy. For Patient 2, the protocol was adapted to the use of a PET-MRI scanner (Biograph mMR) with anatomical and functional MRI.

For both patients, FDG-PET imaging consisted of an injection of 130 MBq of [^18^F]fluoro-deoxy-glucose followed by the acquisition of a static image from 40 to 50 minutes post-injection.

For the activation-task protocol and in accordance with previous studies using fMRI in non-degenerative coma^56^, each patient was asked to mentally visualize himself while sequentially visiting each room in his house. For Patient 1 (PET/CT), the activation-task paradigm was designed as the changes of regional cerebral blood flow (rCBF) at rest and during the visuo-spatial mental imaging. The contrast was estimated by recording the distribution of radioactivity after two repeated intravenous injections of 300 MBq of [^15^O]H2O. For Patient 2 (PET-MR), the acquisition was adapted to an fMRI protocol closely related to the original paradigm^27^ ([45s rest; 45s task] x 10 blocks).

#### Metabolic FDG-PET data analysis

For FDG-PET, a voxel-based analysis was conducted in comparison to a database of 37 healthy subjects^72^ using SPM 12 (Wellcome Department for Cognitive Neuroscience, London, UK) with Matlab R2017a (Mathworks Inc., Natick, MA**^®^**)^72^. Image processing consisted in: i) spatial normalisation to Montreal Neurological Institute (MNI) space for both patients (for Patient 1 in absence of MRI: a temporary explicit mask was defined to allow normalisation by thresholding the patient’s mean uptake value at 2000 Bq/cc to remove relatively high extra-cerebral FDG uptake for the PET data; for Patient 2 the T1 MPRAGE MRI was used for spatial normalisation); ii) common smoothing (kernel size [8 8 8] mm); iii) conversion of PET images to standardized uptake value [SUV] units to reduce weight- and activity-related variability within the database.

Two statistical parametric map analyses were performed at the voxel level to objectively detect global and focal changes of metabolism, applied to each patient separately.

An analysis of variance with factor group (patient vs. healthy controls), age as covariate and proportional scaling to global intracerebral mean SUV was used to assess the global hypometabolism visually observed by the nuclear medicine physician and to search for any voxel with a preserved metabolism in the fronto-parietal associative cortex^56^. Age was defined as co-variate across each patient and the control group to reduce the effect of age-related brain atrophy. A liberal threshold was used for this analysis (uncorrected p < 0.05), according to the methodology proposed by Stender et al^56^.

A second analysis of variance with group factor and co-variables “age” and “global mean value” (extracted from every SUV image within the intracranial volume mask from SPM) was performed to specify the patterns of focal metabolic changes corrected for global activity (*i.e.* relative hypometabolism or relative hypermetabolism in case of global hypometabolism). A more stringent threshold was used for this analysis (uncorrected p < 0.01) to confirm the specificity of local clusters, if any.

#### Activation [^15^O]H2O PET and fMRI data analysis

The analysis was focused on parahippocampal areas, under the assumption that they would be activated by the spatial navigation task^27^.

For patient 1 (PET/CT), the activation results from the paired [^15^O]H2O PET scans were analysed using a SISCOM (Subtraction Ictal SPECT Co-registered to MRI) protocol^73^ to allow for a statistical analysis based on voxel-by-voxel differences between the two single images. For patient 2 (PET-MR), the contrast between BOLD imaging (task – rest) was processed in SPM 12 using a general linear model contrasting periods of active imagery with periods of rest^27^. A p-value of < 0.05 (with family wise error rate – FWE – correction for multiple comparisons) was considered to be significant to reliably demonstrate the presence of an activation pattern.

## Data Availability

All data produced in the present work are contained in the manuscript.

## Abbreviations

Act-Pass: Active-Passive auditory oddball paradigm
ALS: Amyotrophic Lateral Sclerosis
ANCOVA: Analysis of Co-Variance
BCI: Brain Computer Interface
CJD: Creutzfeldt-Jakob Disease
cLIS: complete Locked-In Syndrome
CT: Computerised Tomography
EEG: Electroencephalogram
EP: Evoked Potential
FGD: Fluoro-Deoxy-Glucose
fMRI: functional MRI
FTD: Fronto-Temporal Dementia
MAAB: Multimodal Awareness Assessment Battery
MCS: Minimal Conscious State
MMN: Mismatch Negativity
PET: Positron Emission Tomography
SON: Subject Own Name
SUV: Standardized Uptake Value
SWS: Slow Wave Sleep

---

**<u>Figure 1:</u> Study design in relation to the medical history**

For Patient 1, the first part of the MAAB (Time 1, T1) was performed 5 years after onset and confirmed the feasibility of communicating via BCI. Two further attempts at BCI-based communication were performed at the patient’s bedside 6 years after onset and failed to establish a reliable communication. During the BCI sessions, an abnormally low frequency EEG activity was observed with an inconstant reactivity to nociceptive stimulus. The patient was admitted 2 months later (Time 2, T2) for another MAAB and a complementary imaging evaluation (CT scan, activation-task PET and FDG PET).

For Patient 2, a single MAAB was planned 8 years after onset, together with the attempt of BCI-based communication and the complementary evaluation (MRI scan, activation-task functional MRI and FDG PET).

**<u>Figure 2:</u> Neurophysiological data: Classic analysis, Active - Passive paradigm, and BCI results**

**A – Summary table for neurophysiological results**

**B – Active ERP paradigms**: For the “Count”, the deviant response is shown in red and the standard response in green. For Patient 1 at T1, the comparison between red lines and green lines shows the presence of a statistically significant P3b (grey bars). For the “Focused *vs* Diverted”, the focused-attention response is shown as a continuous red line and the diverted-attention response as a dotted red line. The comparison between continuous and dotted red lines shows the presence of a statistically significant N2 for Patient 1 at T1 and an inconsistent late parietal P3 for Patient 2. Grey shaded bars indicate the latencies presenting statistically significant differences.

SEPs: Somatosensory Evoked Potentials

BAEPs: Brainstem Auditory Evoked Potentials

MLAEPs: Middle Latency Brainstem Auditory Evoked Potentials

**<u>Figure 3:</u> Neurophysiological data: background EEG activity**

Selected results from ASEEGA algorithm (Physip®). On the x-axis: time of the polysomnography. On the y-axis, from top to bottom: time-frequency analysis of EEG (normalized power spectral density in dB), mean frequency of spindles per 30-seconds epoch, 5-class hypnogram (W = Wake, N1-2-3 = Sleep stage 1-2-3, R = REM sleep, Art: Artifacts. Results are presented for a healthy subject then for each patient. An illustration by raw EEG pages is provided for the most relevant patterns.

*Patient 1 at T2:* Abnormal wakefulness pattern with a pathological wake EEG on the left and an N3 period on the right (Delta dominant frequency corresponding to a slow wave sleep period).

On the time-frequency analysis, the increase of Theta-Delta frequency during the prolonged wake period at T2 can be compared to the time-frequency analysis made on a 20-min EEG at T1 (predominant Alpha-Theta frequency without Delta rhythms) and to the pattern of the healthy subject.

*Patient 2*: Abnormal sleep-wake architecture with: i) prolonged periods of wake during the evening (on the left) and the night (around midnight for instance); ii) prolonged sleep periods during the day (N2 period in the middle) and the night (N3 period on the right).

EEG: bipolar montage, pages of 20 seconds, filters = 0.053 – 60 Hz, amplitude = 50 microV/cm.

**<u>Figure 4:</u> Morphological and metabolic imaging**

*Column A:* Patient 1. For PET illustration, the results are projected on the mean PET image of healthy subjects (in the absence of MRI).

*Column B:* Patient 2. For PET illustration, the results are projected the simultaneously acquired MRI.

*Top part:* Focal lobar frontal and temporal atrophy (CT scan for Patient 1) and diffuse atrophy (MRI scan for Patient 2).

*Middle part:* [^18^F]FDG-PET contrast “Patient < Healthy subjects” using an analysis of variance (on SUV images, uncorrected, p<0.05) demonstrating global hypometabolism, in particular for the associative fronto-parietal cortex.

*Bottom part:* [^18^F]FDG -PET contrast “Patient > Healthy subjects” using an analysis of variance (on SUV images, uncorrected, p<0.05) demonstrating global hypermetabolism.

Colour bars: t-scores. All illustrations are shown for the MNI coordinates (X=-1.45, Y=3, Z=-0.68).

**<u>Table 1:</u> Neurophysiological data - BCI paradigm: experimental design and classification results for each patient**

Online and offline results indicating the absence of BCI control by both patients and the high cross-validation accuracy consistent with overfitting.

For p>0.05, chance levels are:

- at 62.5 % for 40 trials
- at 70 % for 20 trials
- at 83 % for 12 trials.

**<u>Figure SOM-1:</u> EEG reactivity during wake period**

*Top:* Patient 1 at T1. Alpha background rhythm with acceleration to the sensory stimulus “touching right arm” (blue line, “on lui touche le bras dt”).

*Bottom:* Patient 2. Mixed frequency background rhythm with acceleration to the auditory stimulus “calling the patient by his name” (blue line, “appel nom prénom”). EEG: bipolar montage, pages of 20 seconds, filters = 0.053 – 60 Hz, amplitude = 50 microV/cm.

**<u>Figure SOM-2:</u> Complement of PET analysis**

*Column A:* Patient 1. For PET illustration, the results are projected on the mean PET image of healthy subjects (in absence of MRI)

*Column B:* Patient 2. For PET illustration, the results are projected the co-acquired MRI.

*Top part:* [^18^F]FDG-PET contrast “Patient < Healthy subjects” using an analysis of variance with 2 co-variables (on SUV images, uncorrected p<0.01) describing relative hypometabolism.

*Bottom part:* [^18^F]F DG-PET contrast “Patient > Healthy subjects” using an analysis of variance with 2 co-variables (on SUV images, uncorrected p<0.01) describing relative hypermetabolism.

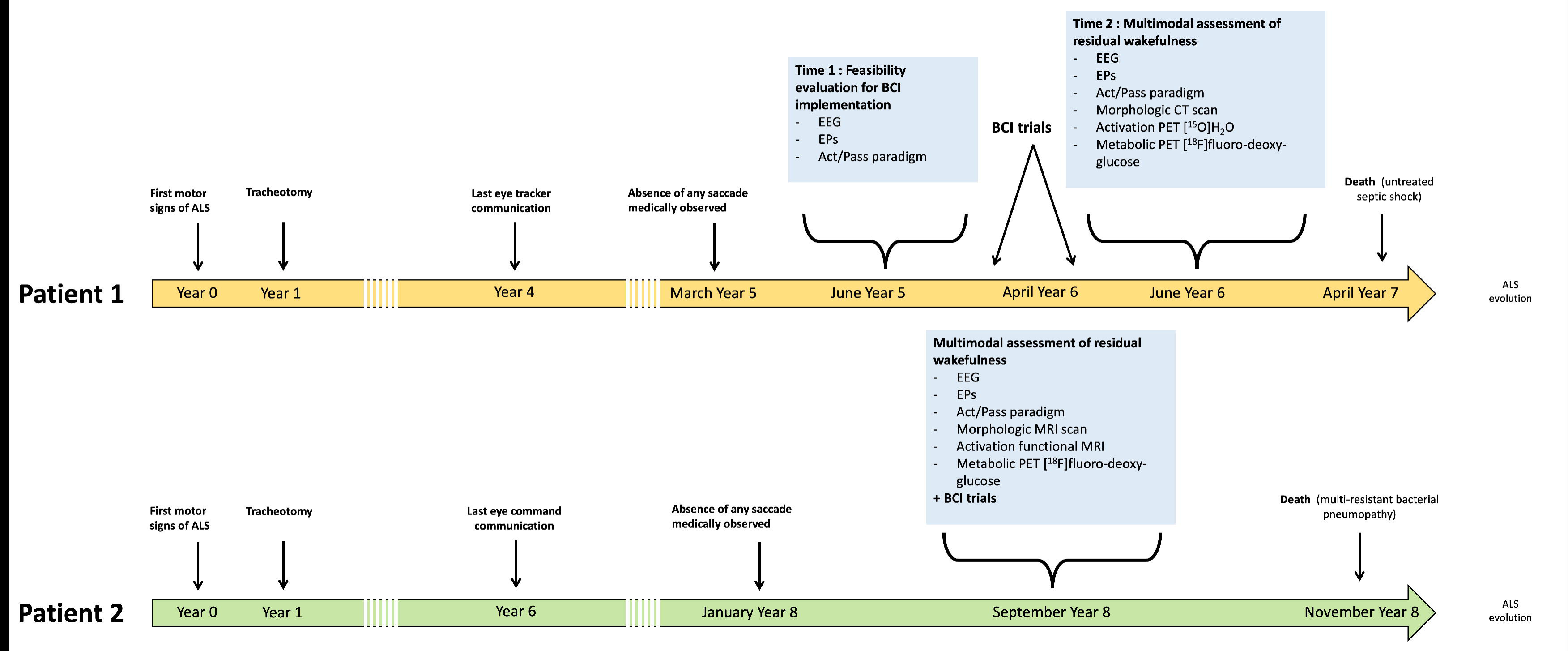

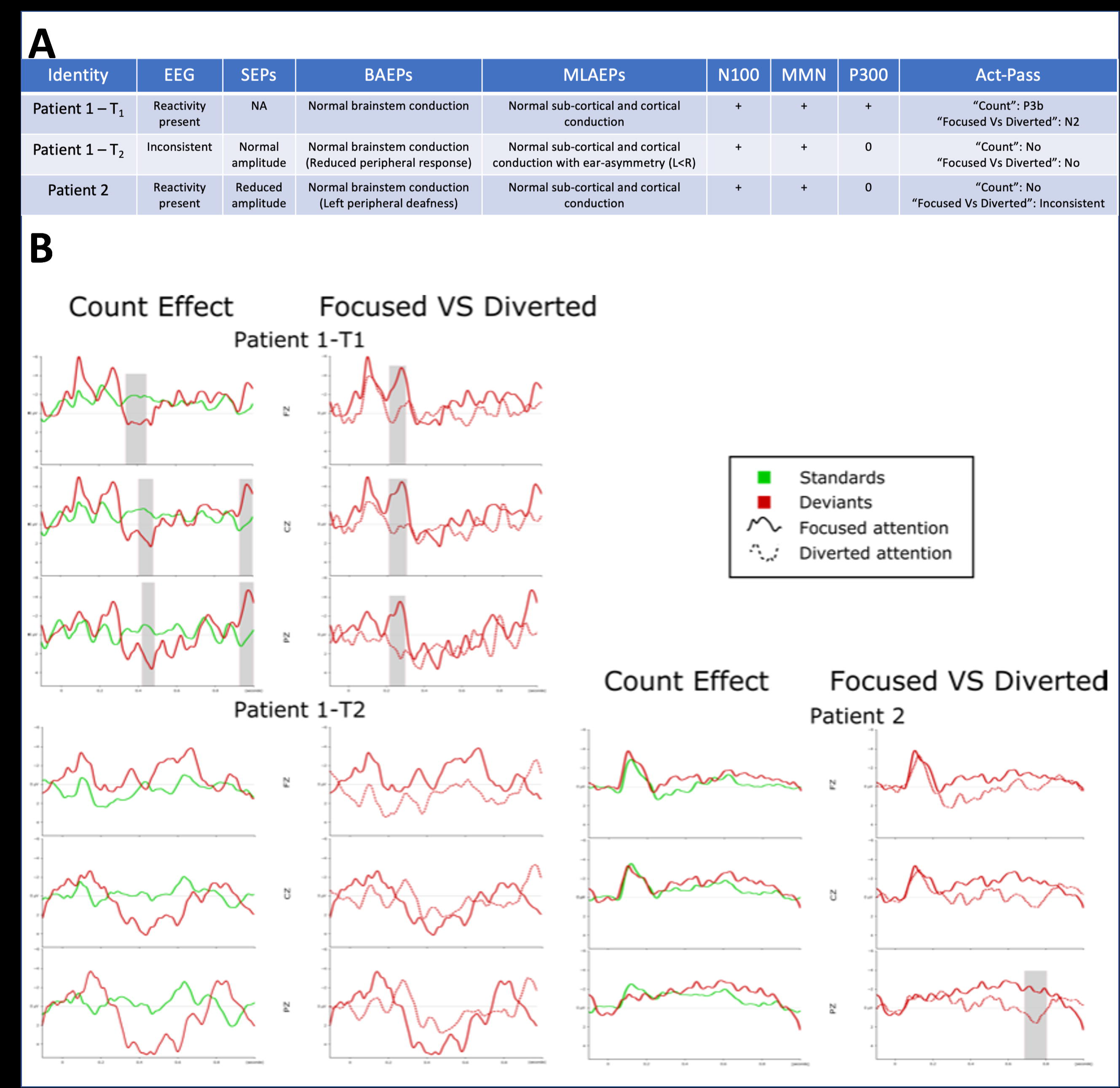

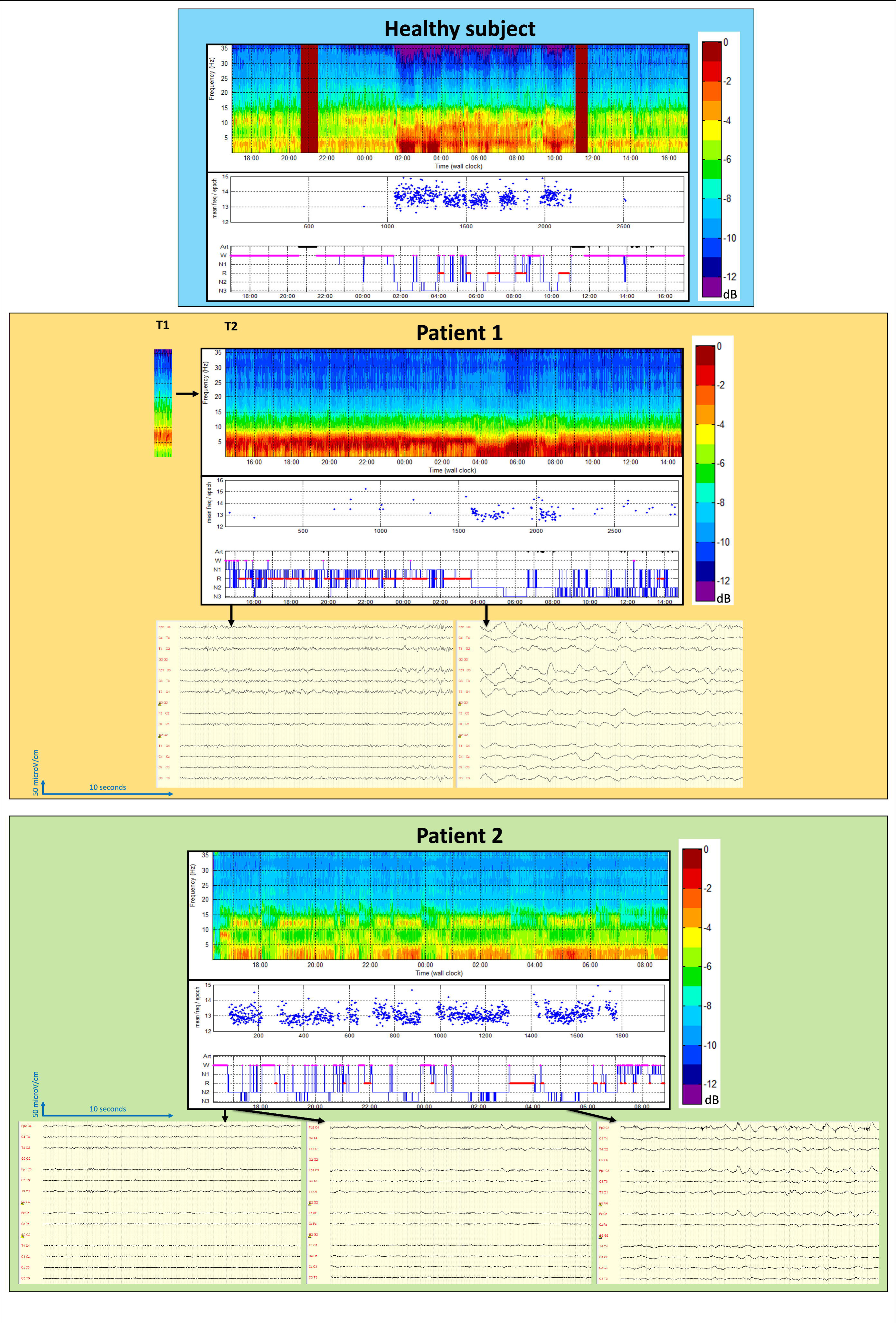

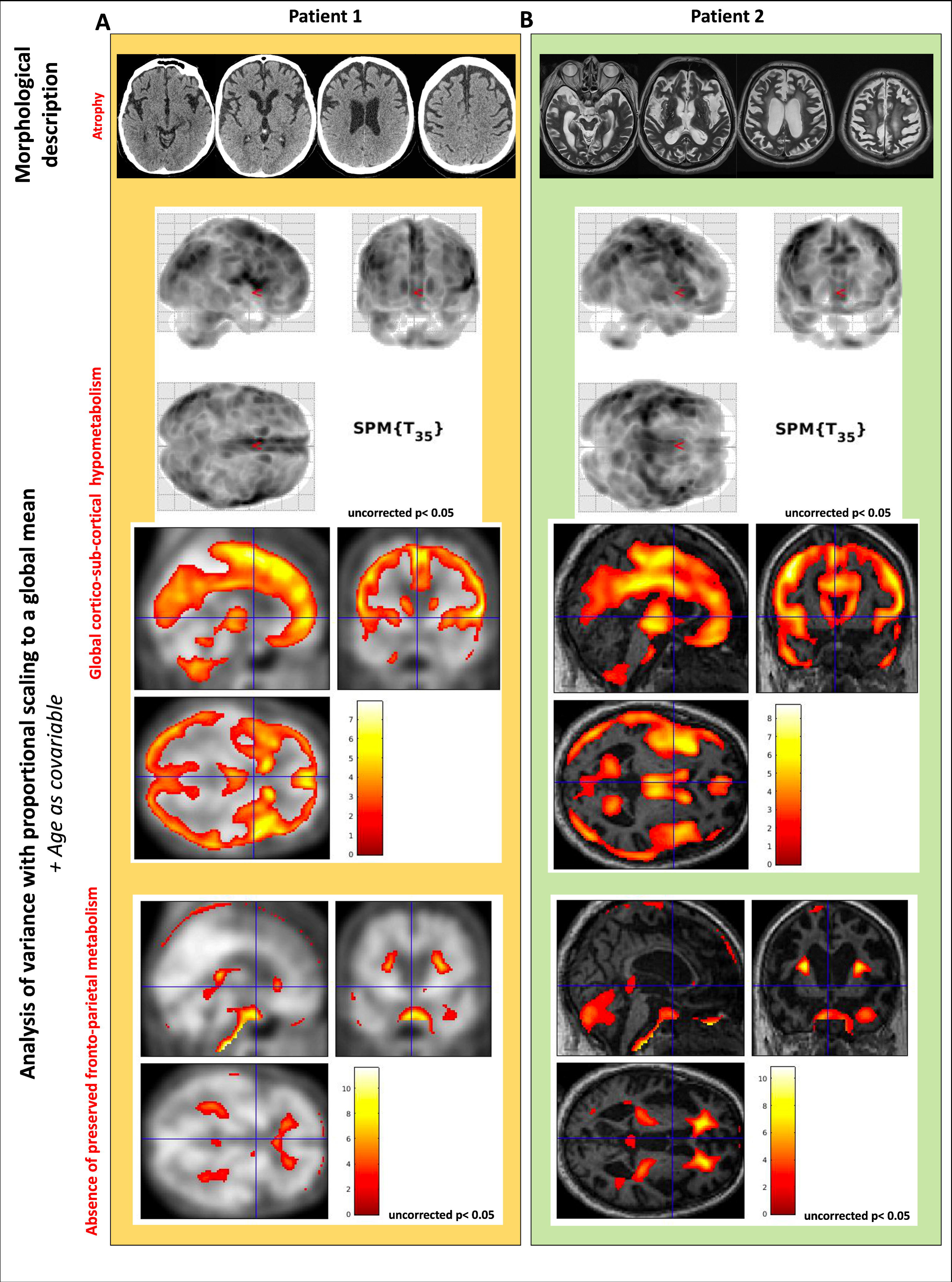

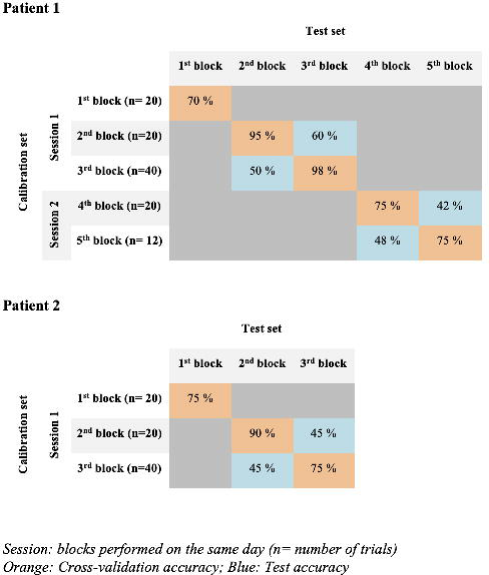

