## Supplementary figures and images for "A disorder of consciousness rather than complete locked-in may be the final stage of ALS"

### Supplementary Figure 1

#### Patient 1, T1 : Presence of reactivity to touch

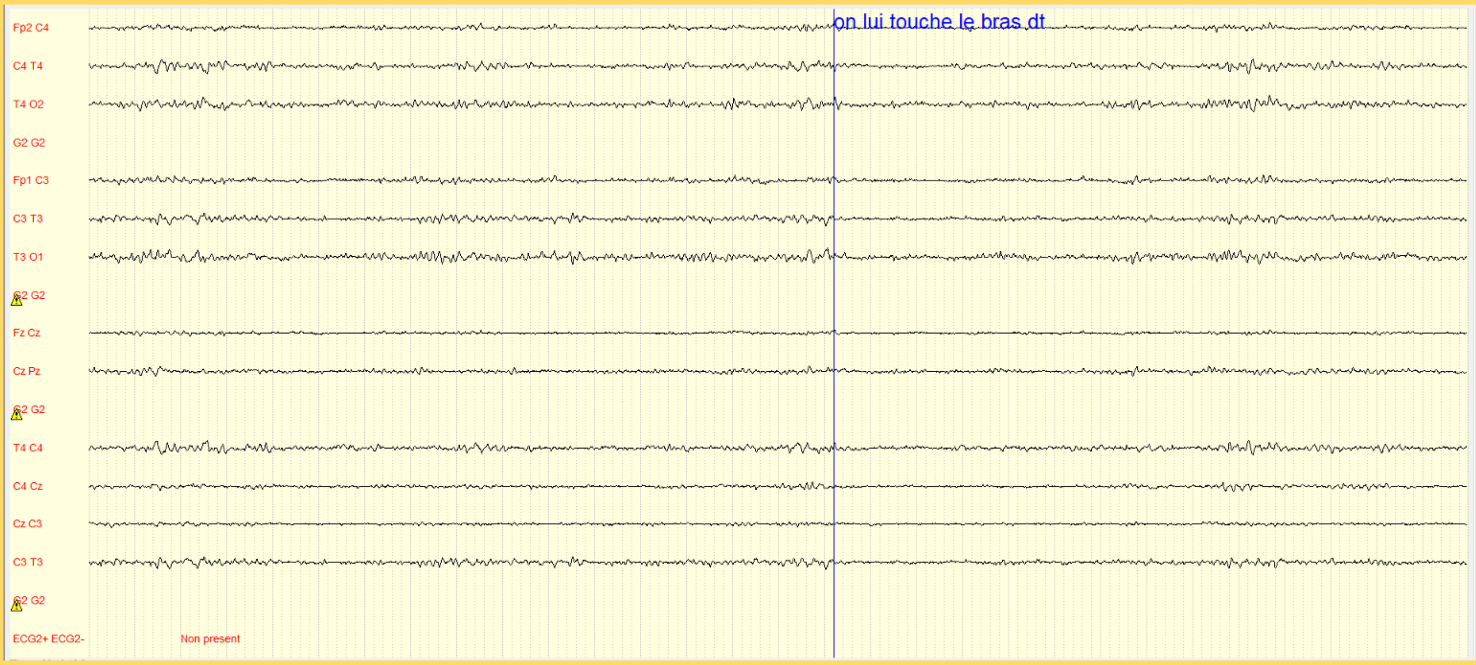


#### Patient 2: Presence of reactivity to sound

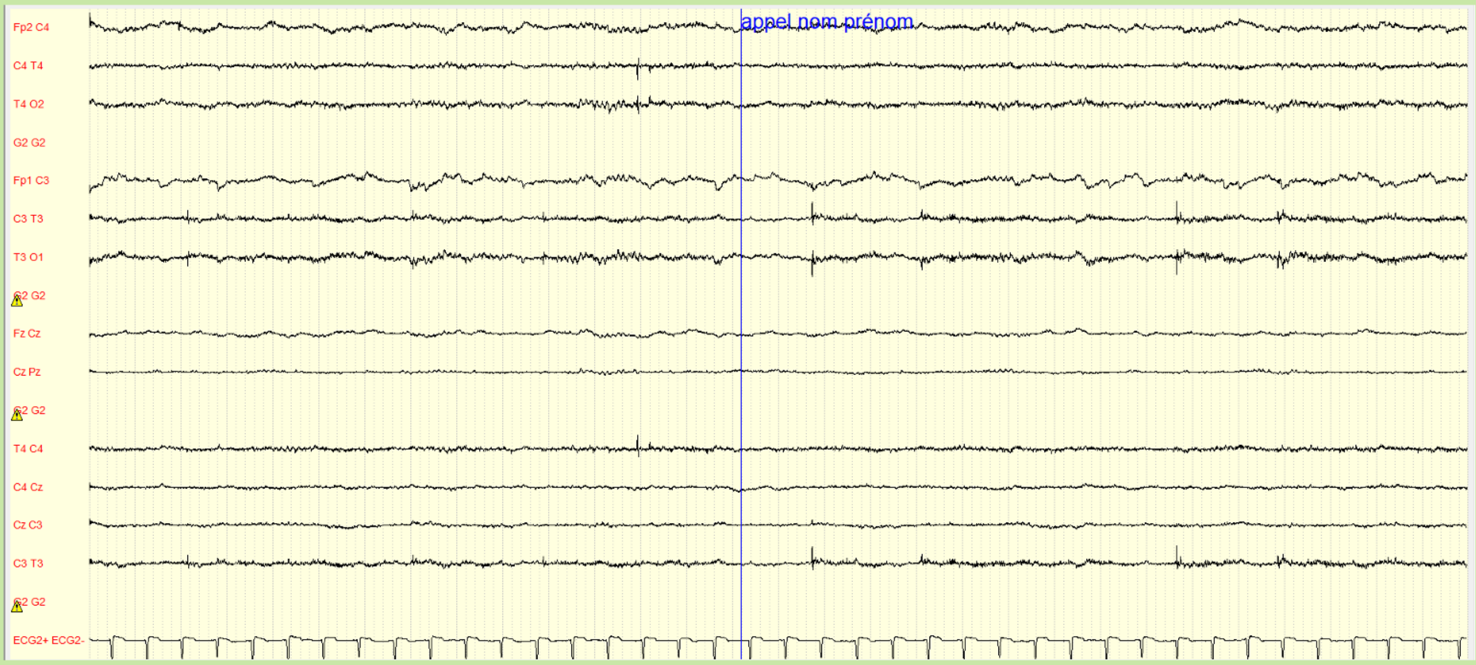

### Supplementary Figure 2

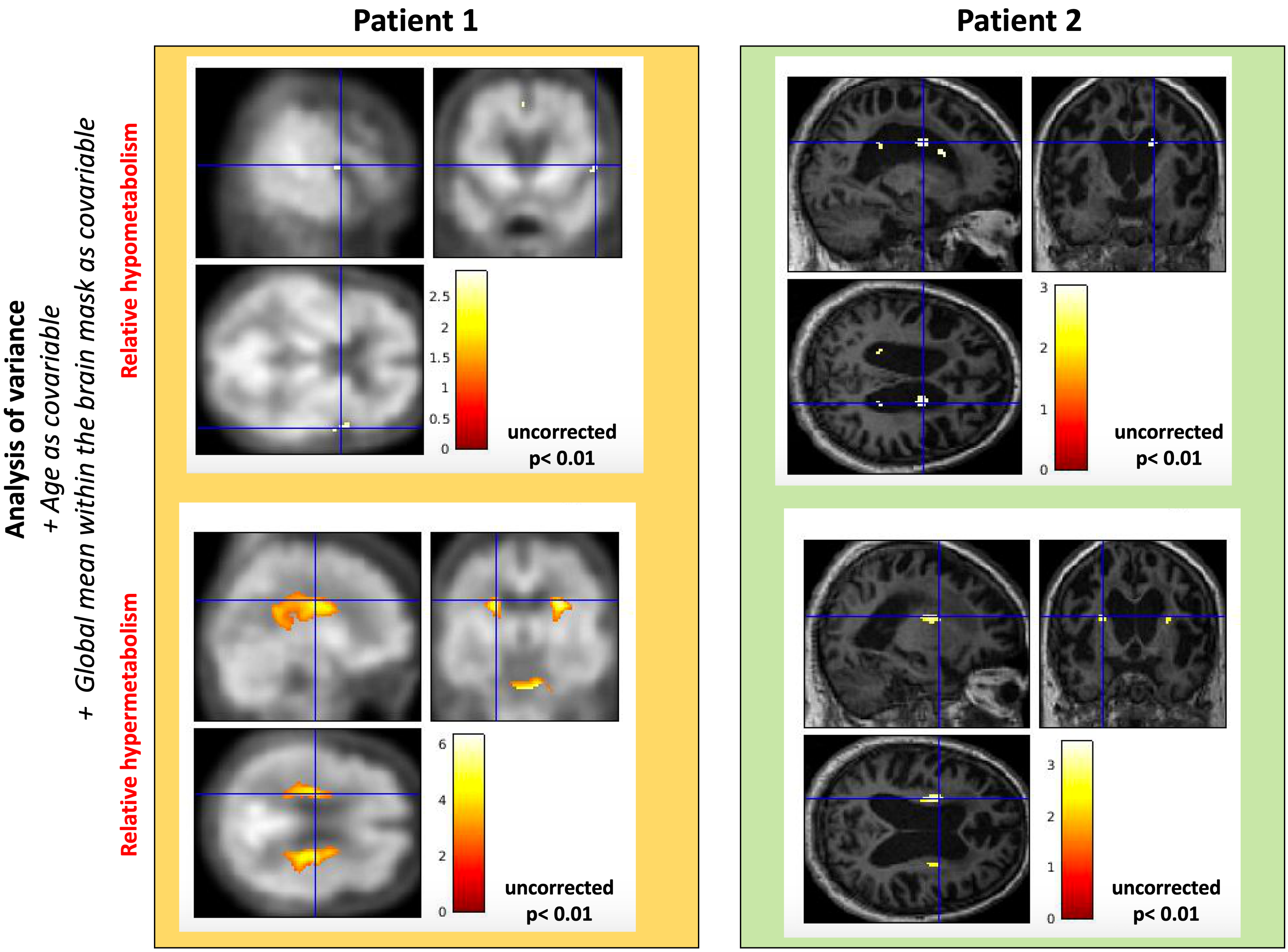
